# Ultrasound measures of intrinsic foot muscle size and motor activation following lateral ankle sprain and chronic ankle instability. A cross-sectional study

**DOI:** 10.1101/2020.07.27.20163220

**Authors:** John J. Fraser, Rachel Koldenhoven, Jay Hertel

## Abstract

**Objectives:** To assess the effects of ankle injury status on intrinsic foot muscle (IFM) size at rest and during contraction in young adults with and without a history of lateral ankle sprain (LAS) and chronic ankle instability (CAI).

**Methods:** Foot Posture Index (FPI), Foot Mobility Magnitude (FMM), and ultrasonographic cross-sectional area of the abductor hallucis (AbdH), flexor digitorum brevis (FDB), quadratus plantae (QP), and flexor hallucis brevis (FHB) were assessed at rest, and during non-resisted and resisted contraction in 22 healthy (13 females, BMI: 22.5±3.2, FPI: 4.2±3.9, FMM: 2.5±1.8), 17 LAS (9 females, BMI: 24.1±3.7, FPI: 2.5±3.4, FMM: 2.7±1.7), 21 Copers (13 females, BMI: 23.7±2.9, FPI: 3.6±4.1, FMM: 1.8±1.3), and 20 CAI (15 females, BMI: 25.1±4.5, FPI: 4.4±3.6., FMM: 2.3±1.1).

**Results:** A multiple linear regression analysis assessing group, sex, BMI, FPI, and FMM on resting and contracted IFM size found sex (*p*<.001), BMI (*p*=.01), FPI (*p*=.05), and FMM*FPI interaction (*p*=.008) accounted for 19% of the variance (*p*=.002) in resting AbdH measures. Sex (*p*<.001) and BMI (*p*=.02) explained 24% of resting FDB measures (*p*<.001). Having a recent LAS (*p*=.03) and FMM (*p*=.02) predicted 11% of non-resisted QP contraction measures (*p*=.04), with sex (*p*<.001) explaining 13% of resting QP measures (*p*=.02). Both sex (*p*=.01) and FMM (*p*=.03) predicted 16% of resting FDB measures (*p*=.01). There were no other statistically significant findings.

**Conclusions:** IFM resting ultrasound measures were primarily determined by sex, BMI, and foot phenotype and not injury status. The clinical utility of these IFM ultrasonographic assessments in young adults with LAS and CAI may be limited.

**Disclosures:** This study was funded in part by the University of Virginia’s Curry School of Education Foundation and the Navy Medicine Professional Development Center, US Navy Bureau of Medicine and Surgery. The views expressed in this manuscript reflect the results of research conducted by the author(s) and do not necessarily reflect the official policy or position of the Department of the Navy, Department of Defense, nor the U.S. Government. Lieutenant Commander John J. Fraser is a military service member and this work was prepared as part of his official duties. Title 17, USC, §105 provides that ‘Copyright protection under this title is not available for any work of the U.S. Government.’ Title 17, USC, §101 defines a U.S. Government work as a work prepared by a military service member or employee of the U.S. Government as part of that person’s official duties. All participants provided informed consent and this study was approved by the Institutional Review Board for Health Sciences Research at the University of Virginia in compliance with all applicable Federal regulations governing the protection of human subjects.

## INTRODUCTION

Lateral ankle sprains (LAS) are ubiquitous, with more than 2-million individuals in the general public affected in the United States annually.^1^ While more than half of individuals who incur a LAS have resolution of symptoms and resumption of physical activity within the first year (Copers),^2^ many will have residual symptoms and persistent activity limitation. Forty percent of individuals who incur a substantial LAS will progress to develop Chronic Ankle Instability (CAI),^3^ a condition characterized by persistent perceived or episodic ‘giving way’ of the ankle that affects function beyond one year following an index injury. Functional limitation in this clinical population is multifactorial and may include varying degrees of mechanical and neurophysiological deficit,^4^ including impairment in the foot.^5,6^

Connective, contractile, and neural impairment in the ankle-foot complex contributes to activity limitation and participation restriction following LAS.^5,6^ Tibial nerve impairment is common following LAS, with up to 83% of individuals found to have reduced nerve conduction following injury.^7,8^ This is clinically meaningful since the extrinsic muscles originating in the superficial and deep posterior compartment and the plantar intrinsic foot muscles (IFM) are innervated by the tibial nerve. Large and significant reductions in plantarflexion, hallux flexion, and lesser toe flexion strength have been observed in individuals with recent LAS and CAI.^5^ While toe flexion is actuated in part by the long toe flexors originating in the leg, these measures have been found to be good surrogates for IFM neuromotor function.^9^ The previously reported findings in individuals with LAS and CAI may indicate, in part, deficit in the IFM.^5^ This supposition is further supported by evidence of reduced IFM volume measured with Magnetic Resonance Imaging (MRI) in young adults with CAI.^10^

While MRI may help clinicians to elucidate the etiology of impairment and provide insight regarding changes in muscle morphology over time, limited access and the high cost associated with this modality likely precludes widespread clinical use following LAS.^11^ Furthermore, MRI performed at a single time point cannot provide information regarding neuromotor function at the point of care.^6,11^

Electromyography may also be a feasible modality to provide insight regarding IFM neuromotor function. However, muscle cross-talk, lack of access to equipment, and relatively few clinicians trained in electrophysiological assessment preclude regular use in the clinical setting.^11^ Ultrasound imaging (USI) is an ubiquitous clinical modality that may have clinical utility at the point of care for assessment of real-time IFM neuromotor function and morphologic changes over time.^6,11^ Serial assessments of resting IFM measures over time may provide insight regarding changes in muscle size resulting from neuromotor impairment or in response to exercises.^6,11,12^ Assessments of non-resisted or resisted IFM activation may provide insight to neuromotor function at the point of care.^6,11,12^ While these measure have been found to have excellent measurement properties,^11^ the clinical utility of USI of the IFM in the assessment of patients following LAS and CAI is unclear. Furthermore, IFM size maybe influenced by sex, body mass, and foot phenotype,^9,13^ factors that will need to be considered in this clinical population. Therefore, the purpose of this cross-sectional study was to assess the effects of ankle injury status on IFM size at rest and during contraction in recreationally-active young adults with LAS, Copers, CAI, and healthy controls, while factoring the influences of sex, body mass index, and foot phenotype.

## METHODS

### Design

A laboratory-based, descriptive cross-sectional study was performed. The independent variable was group (Control, Coper, LAS, CAI) and the dependent variables were IFM cross-sectional area at rest and motor activation ratios during non-resisted and resisted contraction. The relationships between IFM resting size, motor activation, and clinical measures of clinical measures of foot morphology, posture, and strength were also assessed.

### Participants

A convenience sample of 80 recreationally-active individuals (Control: n=22, Coper: n=21, LAS: n=17, CAI: n=20) aged 18-35 with and without history of a LAS and CAI were recruited from a public university for participation from September 2016 to May 2017. The participants in this study were part of larger study of multisegmented foot function, with demographic and injury history have been previously reported.^5^ **Table 1** details group demographic, injury history, and patient-reported outcome measures. There were no significant group differences in age, anthropometrics, or foot phenotype. “Recreationally-active” was defined as participation in some form of physical activity for at least 20-minutes per day, at least thrice weekly. Participants were included in the Control group if they did not have any history of ankle or foot sprain. Individuals who sustained an inversion sprain that affected function 2-8 weeks prior to consent were included in the LAS group. Participants with a history of at least one LAS at a minimum of 12-months prior to the study who did not experience perceived or episodic giving way and scored Identification of Functional Ankle Instability (IdFAI) ≤10, Foot and Ankle Ability Measure (FAAM) activities of daily living subscale (ADL) ≥99 and FAAM-Sport ≥97 were included in the Coper group.^2^ Individuals with a history of at least one LAS at a minimum of 12-months prior to the study who experienced continued perceived or episodes of giving way and scored IdFAI >10, FAAM-ADL <90 and FAAM-Sport < 85 and did not sprain their ankle in the past 8-weeks were included in the CAI group.^14^ Individuals were excluded if they had any self-reported history of fracture or surgery in the leg or foot, self-reported disability due to neuromuscular impairment in the lower extremity, neurological or vestibular impairment that affected balance, diabetes mellitus, lumbosacral radiculopathy, a soft tissue disorder such as Marfan or Ehlers-Danlos syndrome, any absolute contraindication to ankle or foot joint manipulation, or were pregnant. Participants who met inclusion criteria provided informed consent. Recruitment was stratified to ensure relative equity in the distribution of males and females in each group. **Figure 1** details the study flow sheet from recruitment to analysis. Data was collected in the university’s sports medicine laboratory. The study was approved by a university’s Institutional Review Board.

**Table 1.**
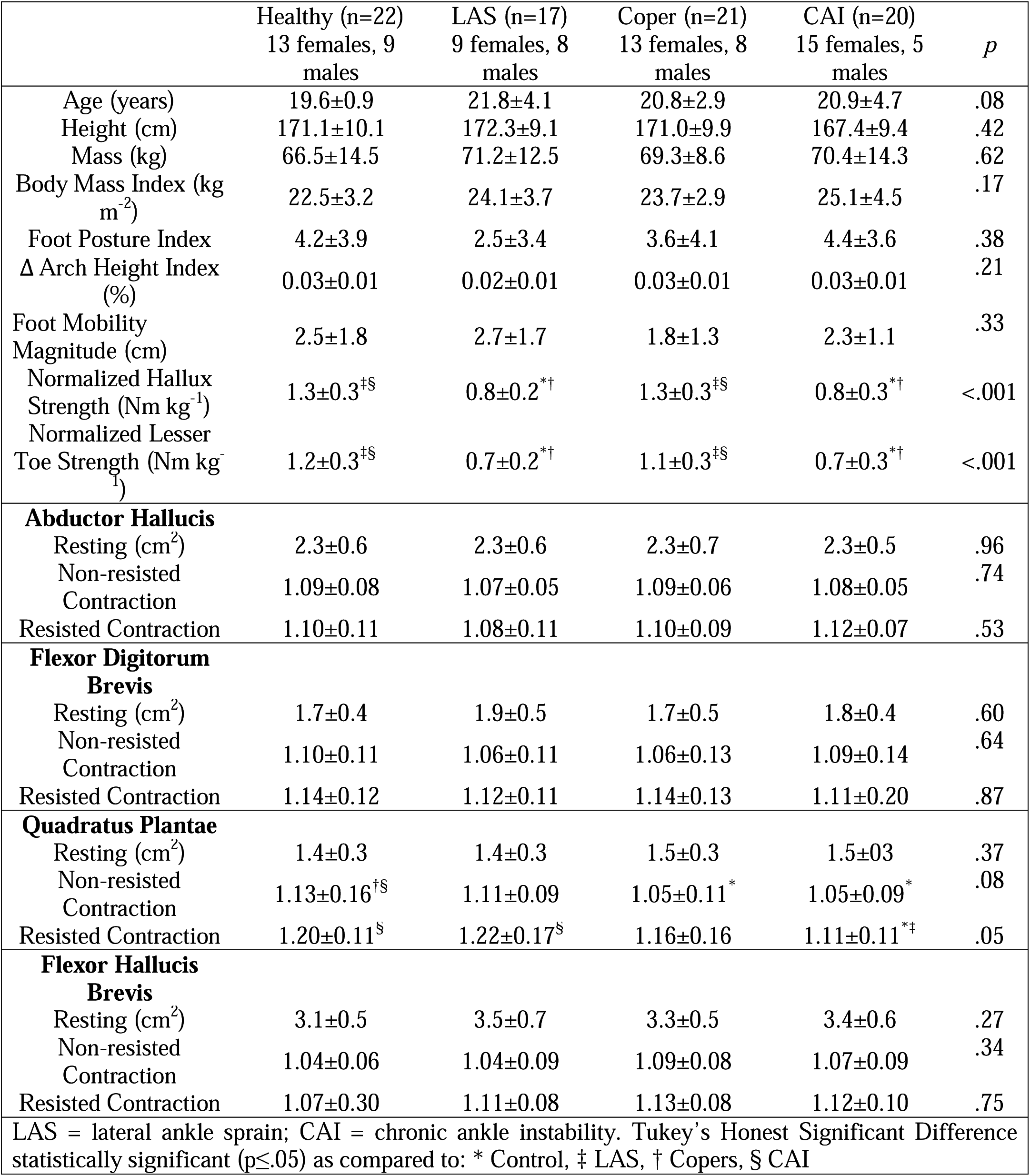
Participant demographic and ultrasound measures.

**Figure 1.**
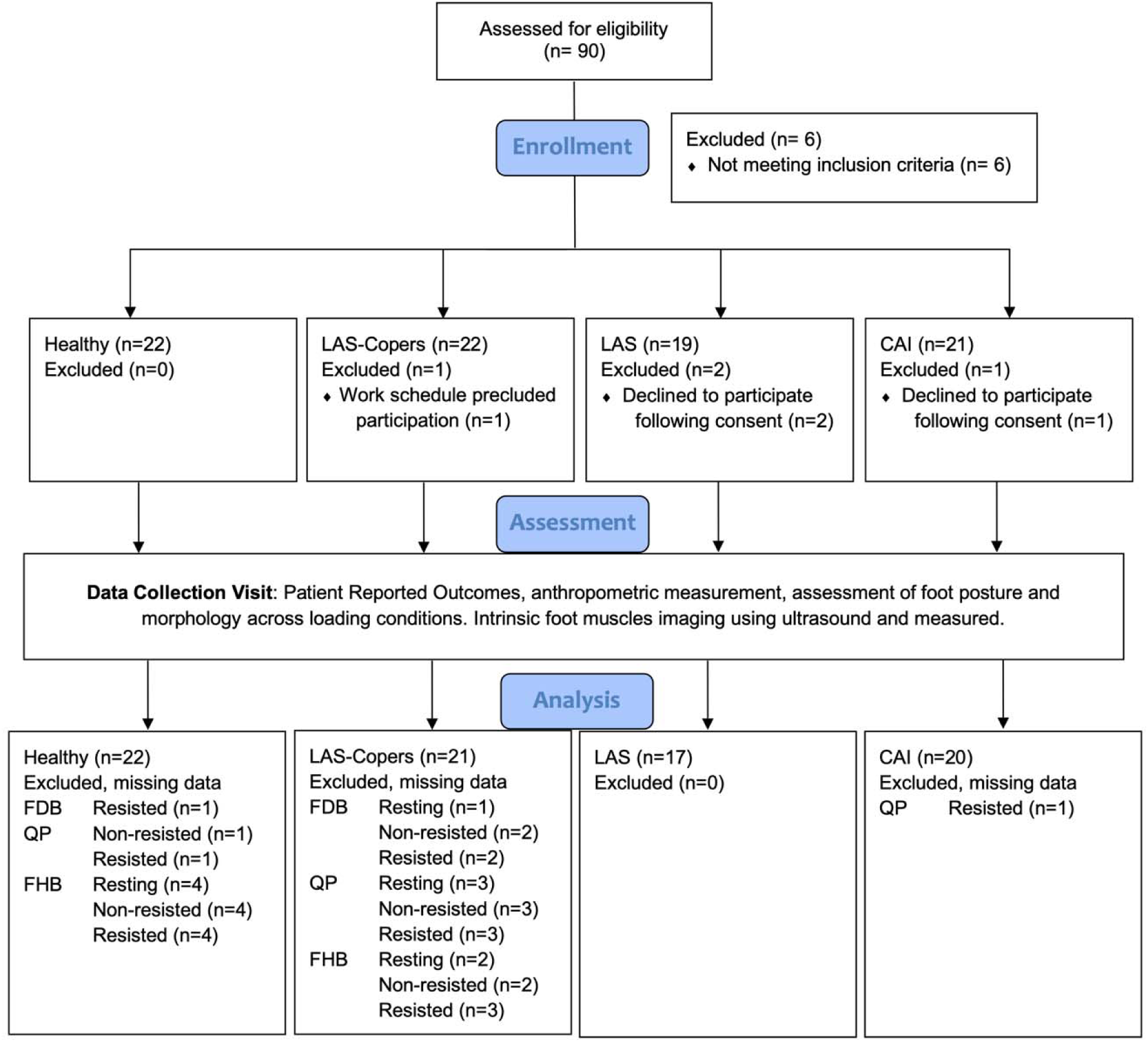
Study flow sheet from recruitment to analysis. LAS, Lateral ankle sprain; CAI. Chronic Ankle Instability; FDB, flexor digitorum brevis; QP, quadratus plantae; FHB, flexor hallucis brevis.

### Procedures

Following consent, participants provided demographic information, health and injury history, and completed the FAAM ADL^15^ and Sport subscales,^16^ Identification of Functional Ankle Instability (IdFAI),^17^ the Patient Reported Outcomes Measurement Information System (PROMIS) General Health Questionnaire,^18^ the 11-item Tampa Scale of Kinesiophobia (TSK-11),^19^ and the Godin Leisure-time Exercise Questionnaire.^20^ Predicted EUROQOL (EQ-5D) quality of life scores were calculated using methods described by Revicki and colleagues.^21^ Height, mass, leg length, and foot morphology were measured and foot posture index assessed.^22^

Demographic information, medical history, physical examination of the non-clinical group, and ultrasound imaging were performed by the primary author who was a physical therapist and was a board-certified orthopaedic clinical specialist with 15-years of clinical experience and one-year experience performing USI. Physical examinations of the LAS and CAI groups were performed by either an athletic trainer with three-years clinical experience or a physical therapist with two-years clinical experience who were blinded to the participants’ medical history and functional status. USI measurement was performed by two laboratory assistants familiar with the IFM anatomy and were blinded to the participant’s clinical history and state of contraction. In the LAS and CAI groups, the involved limb was assessed. In the case of participants with a history of bilateral injury, the self-reported worse limb was examined. The limbs of healthy controls were matched to the limbs assessed in the clinical groups.

#### Morphologic Foot Assessment

Foot posture was assessed in standing using the Foot Posture Index–6 item version (FPI), a categorical measure of foot type that is based on five observations and one palpatory assessment.^22^ Morphologic measurements were performed using the Arch Height Index Measurement System (JAKTOOL Corporation, Cranberry, NJ). Total and truncated foot length, arch height, and foot width were measured in sitting and standing. Change in arch height index (ΔAHI),^23^ and foot mobility magnitude (FMM)^24^ were calculated from the morphologic foot measurements across loading.

#### Muscle Strength

Muscle strength of hallux flexion, and lesser toe flexion were assessed with the microFET2 digital handheld dynamometer (Hoggan Health Industries, West Jordan, UT) using previously described methods.^25^ An estimate of torque was derived from the product of force and segment length, normalized to body mass, and reported in Newton-meters per kilogram (Nm kg^-1^). For toe flexion strength measures, the ankle was positioned in 45° plantarflexion to reduce contribution of the extrinsic foot muscles through active insufficiency and increase demand of the IFMs.^26^ Measures of hallux and lesser toe strength were normalized to hallux length (difference in total and truncated foot lengths). Strength measures were based on highest force produced among the three trials. In the case of an invalid trial (due to equipment difficulty, deviation from test position, or substitution motion), the participant rested prior to retesting to mitigate effects from fatigue.

#### IFM Muscle Size and Motor Function

USI of the abductor hallucis (AbdH), flexor digitorum brevis (FDB), quadratus plantae (QP), and flexor hallucis brevis (FHB) cross-sectional area during rest, contraction without external resistance at the end range of motion (non-resisted contraction), and maximum voluntary contraction against isometric manual resistance (resisted contraction) were performed as described by Fraser and colleagues.^11^ **Figure 2** depicts the transducer location and measured images of the IFM. USI was performed using a Siemens Acuson Freestyle US system with a wireless 8-Mhz linear transducer (Siemens, Mountain View, CA). Each muscle was imaged thrice at rest, during active contraction, and with resistance. Images were measured by two trained assessors, who were blinded to injury status and physical examination findings, using ImageJ version 1.50f (National Institutes of Health, Bethesda, MD). The average measurement of the three images were utilized for analysis in each condition. Measures of active and resisted contraction were normalized to resting measures and reported as an activation ratio,^11,12,27^ calculated using the formula:

**Figure 2.**
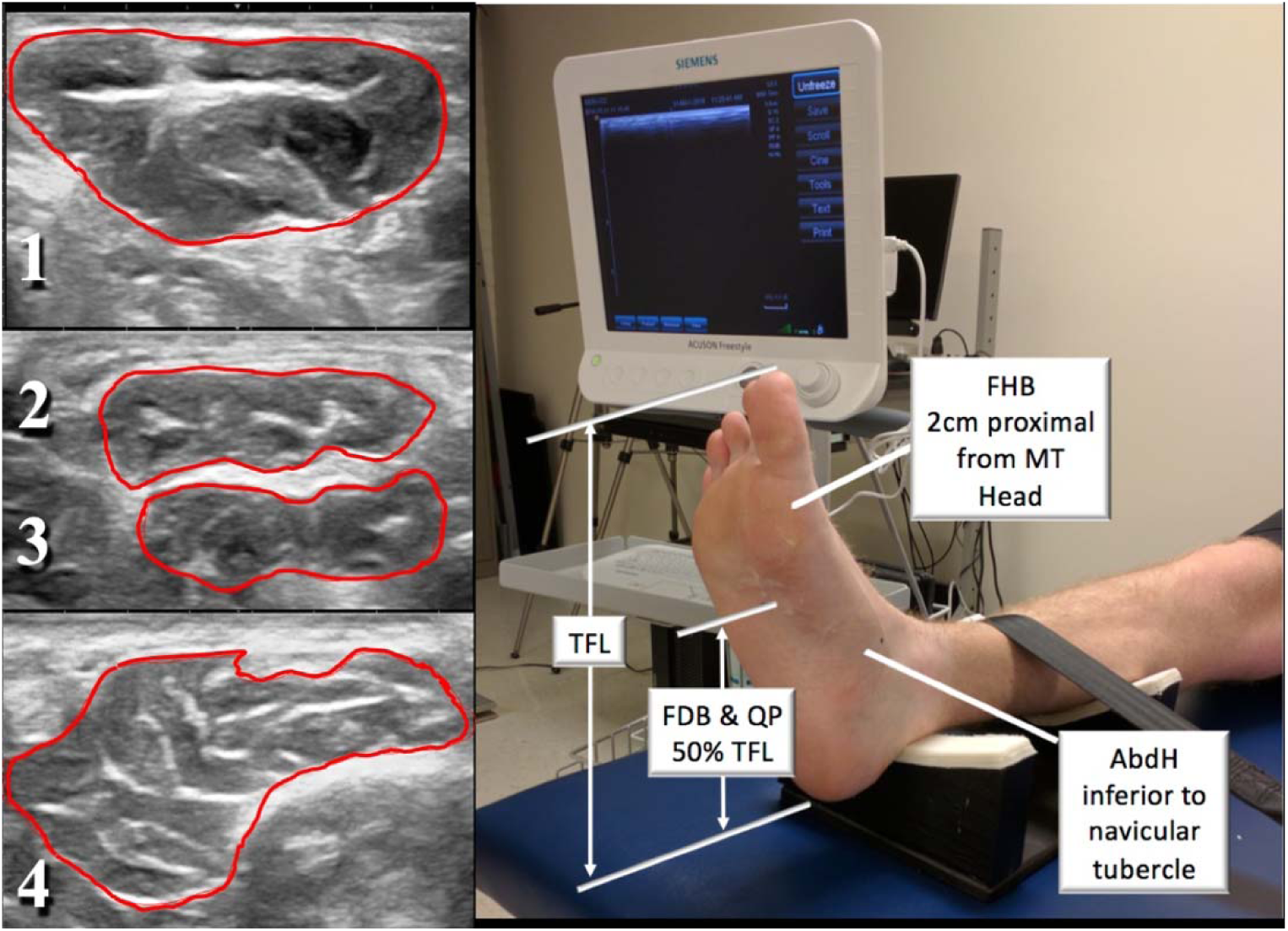
Site of transducer placement during ultrasound scans of the 1. abductor hallucis (Abd. H.), 2. flexor digitorum brevis (FDB), 3. quadratus plantae (QP), and 4. flexor hallucis brevis (FHB). MT, metatarsal; TFL, total foot length. Adapted from Fraser JJ, Mangum LC, Hertel J. Test-retest reliability of ultrasound measures of intrinsic foot motor function. *Phys Ther Sport*. 2018;30:39-47. doi:10.1016/j.ptsp.2017.11.032. Used with permission of the publisher.

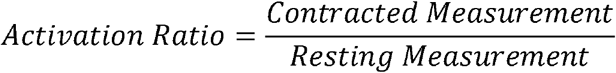

An activation ratio > 1.00 indicates an increase in muscle size and < 1.00 indicative of a decrease in muscle size in comparison to the muscle size at rest.

### Statistical Analysis

The level of significance was *p* ≤0.05 for all analyses. *A priori* sample size estimation of 10 participants per group were needed to demonstrate large effects based on the minimal detectable change in IFM activation ratio of 0.21, a=.05, and =.20.^11^ Descriptive statistics were calculated for group demographic and clinical outcome measures. Ultrasound images that were of poor quality were omitted from analysis.

A preliminary analysis was conducted to assess the effects of group alone on IFM size and activation, without consideration of potential mediating factors. Prior to group comparison analysis, Levene’s test of homogeneity of variance were performed. Variables that had significant homogeneity were assessed using a one-way analysis of variance (ANOVA). Variables that failed to achieve significance with Levene’s test were assessed using Welch’s ANOVA. For significant findings, *post hoc* Tukey’s Honestly Significant Differences and Cohen’s *d* effect size (ES) estimates were calculated for each clinical group compared to healthy controls. ES were interpreted using the scheme proposed by Cohen:^28^ <0.2 equates to a trivial ES, 0.2-0.49 small, 0.5-0.79 moderate, and >0.8 large. Comparisons where the 95% CI around ES point estimate did not cross the ‘0’ threshold were considered statistically significant.

Correlation analysis comparing group, sex, anthropometric, foot composite variables to IFM size at rest and during contraction was performed to inform the development of a regression model. Associations between hallux and lesser toe flexion normalized strength variables and IFM were assessed to evaluate the potential utility of these measures as clinical surrogates. Relationships between continuous variables were evaluated using Pearson’s correlation coefficients. Assessment of relationships with categorical variables (group, sex) were performed using Spearman’s rho. Correlations were interpreted ≥.75 as good to excellent, .50-74 as moderate to good, .25-.49 as fair, and 0-.25 as little to no association.^29^

To assess the contribution of the factors on resting, non-resisted contraction, and resisted contraction measures, three multiple linear regression analyses were performed for each IFM:

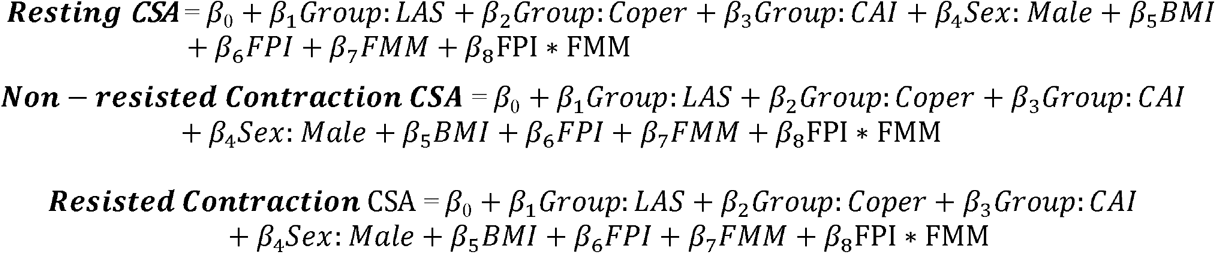

Data was analyzed using R Version 3.5.1 (The R Foundation for Statistical Computing, Vienna, Austria). Cohen’s *d* effect sizes, proportion estimates, and 95% confidence intervals (CI) were calculated using Excel for Mac 2016 (Microsoft Corp., Redmond, WA).

## RESULTS

In the preliminary analysis assessing between-group differences in IFM resting size and activation without consideration of mediating factors (**Table 1**), there were significant group differences observed in the QP non-resisted measures, with the Coper (*d*: 0.58; 95% CI: 0.07-1.09) and CAI (*d*: 0.61; 95% CI: 0.09-1.13) groups demonstrating significant, moderately lower activation compared to healthy controls. Similarly, the CAI group also demonstrated significantly lower activation in the QP resisted measures (*d*: 0.82; 95% CI: 0.29-1.35) compared to both LAS and control groups. **Figure 3** details the assessment of effect sizes comparing LAS, LAS Coper, and CAI groups compared to healthy controls.

**Figure 3.**
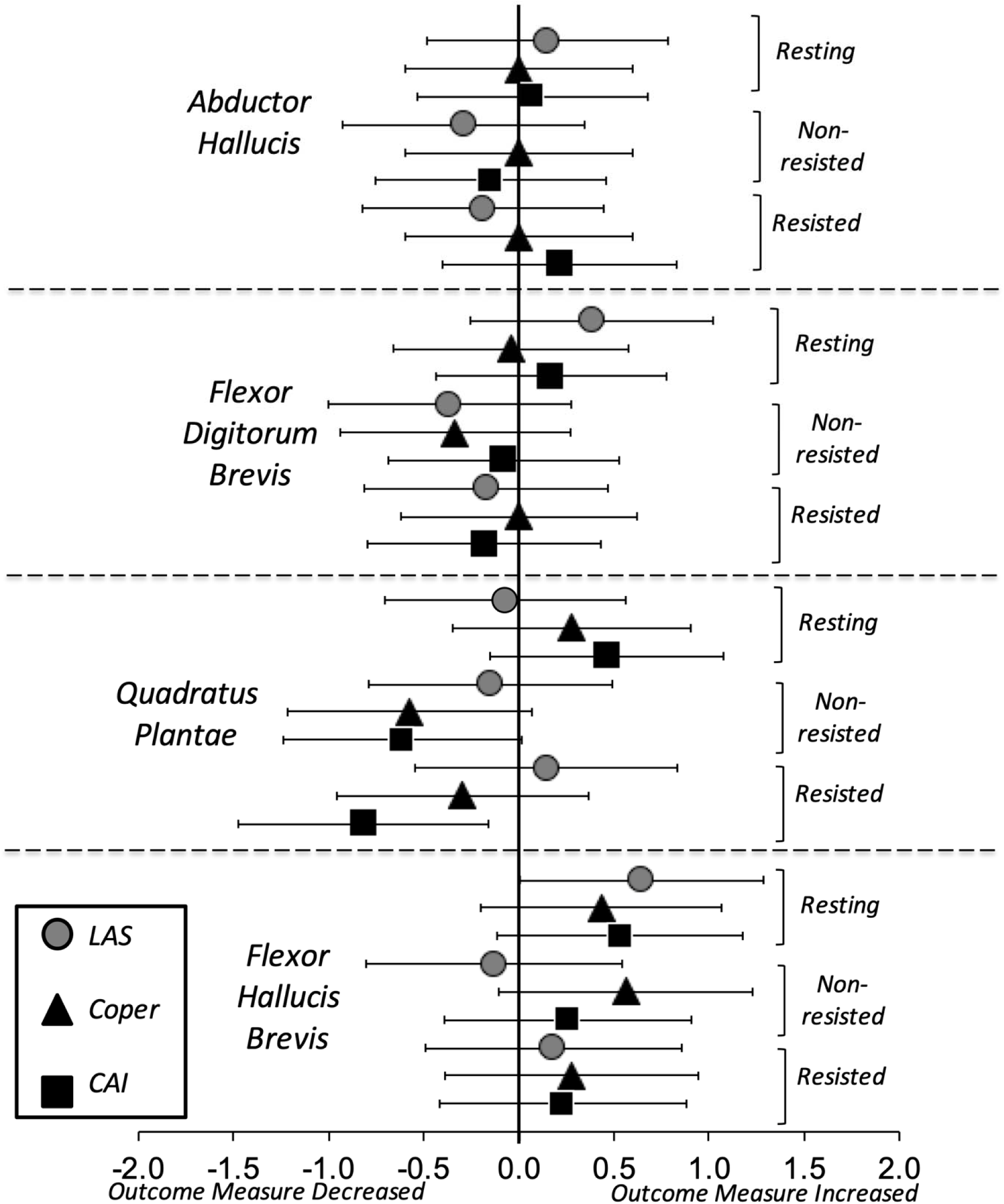
Cohen’s *d* effect size estimates comparing lateral ankle sprain (LAS), LAS Coper, and Chronic Ankle Instability (CAI) groups to healthy controls.

The results of the correlation analysis of participant group, sex, body anthropometrics, morphologic foot measurements on IFM measures is reported in **Table 2**. Group had significant fair associations with non-resisted and resisted measures of the QP. Sex also had significant fair association with resting measures of all four IFMs evaluated. BMI had little to no association with resting FDB measures. Among foot phenotype measures, FMM had fair association with resting FHB and non-resisted QP measures, with ΔAHI demonstrating little to no association with resting FHB measures. Both normalized hallux and lesser toe strength measures had significant, but little to no association with FHB resisted contraction measured with USI.

**Table 2:**
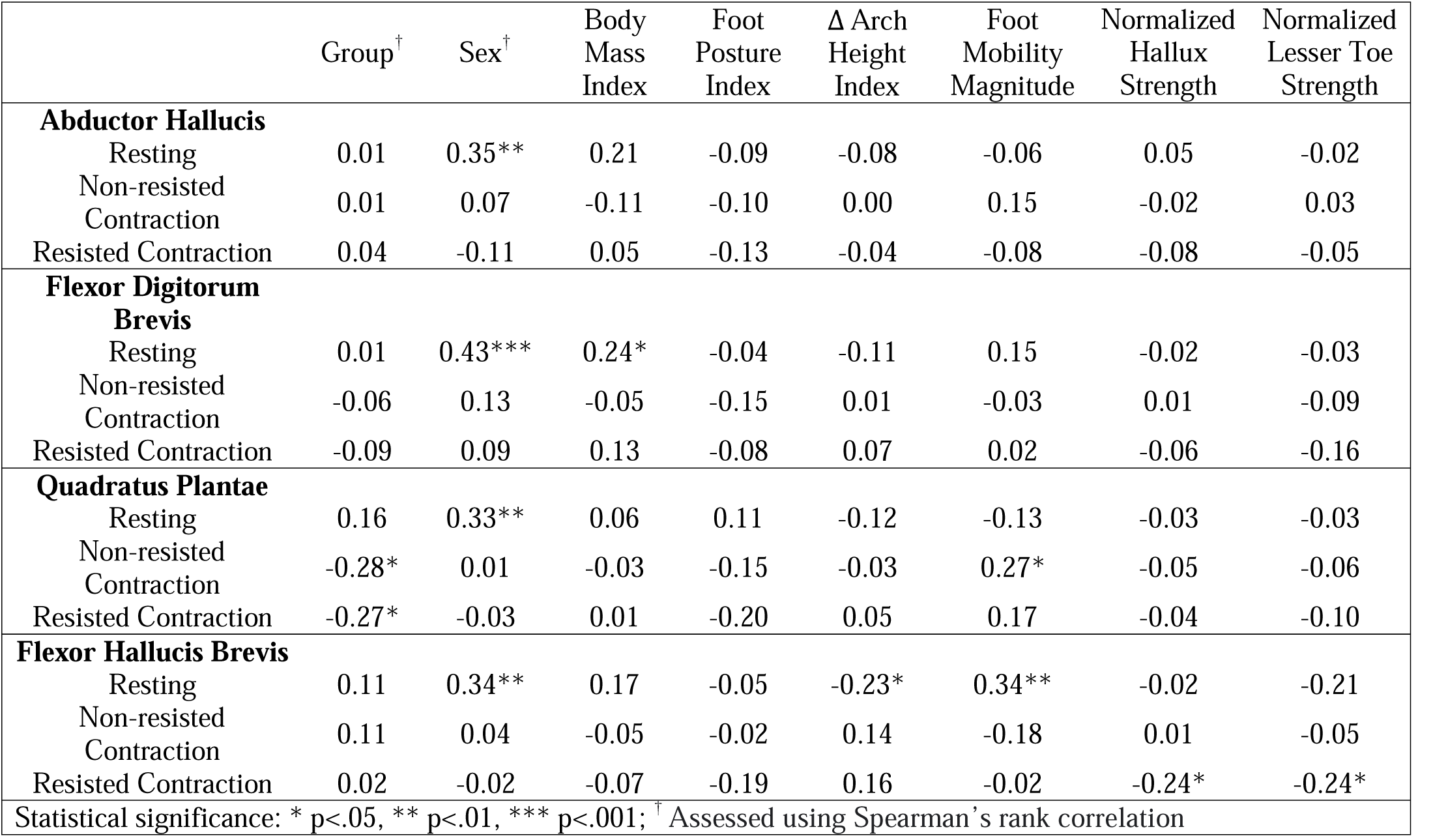
Associations of participant group, sex, body anthropometrics, morphologic foot measurements with ultrasound measures.

The results of the multiple linear regressions are reported in **Tables 3-6**. Sex (*p*<.001), BMI (*p*=.01), FPI (*p*=.05), and FMM*FPI interaction (*p*=.008) predicted 19% of the variance (*p*=.002) in resting AbdH measures. Sex (*p*<.001) and BMI (*p*=.02) explained 24% of resting FDB measures (*p*<.001). Having a recent LAS (*p*=.03) and FMM (*p*=.02) predicted 11% of non-resisted QP contraction measures (*p*=.04), with sex (*p*<.001) explaining 13% of resting QP measures (*p*=.02). Both sex (*p*=.01) and FMM (*p*=.03) predicted 16% of resting FDB measures (*p*=.01). There were no other statistically significant findings.

**Table 3.** Results of the multiple linear regression analysis predicting ultrasound measures of abductor hallucis resting size and during motor activation

|  | Resting |  |  | Non-resisted Contraction |  |  | Resisted Contraction |  |  |
| --- | --- | --- | --- | --- | --- | --- | --- | --- | --- |
|  | B | SE | p | B | SE | p | B | SE | p |
| Constant | 0.92 | 0.44 | .04* | 1.12 | 0.05 | <.001* | 1.09 | 0.00 | <.001* |
| Group: LAS | -0.01 | 0.12 | .95 | 0.01 | 0.02 | .68 | 0.01 | 0.02 | .65 |
| Group: Coper | 0.03 | 0.12 | .77 | 0.01 | 0.01 | .66 | 0.03 | 0.02 | .22 |
| Group: CAI | 0.02 | 0.13 | .87 | -0.02 | 0.02 | .22 | -0.01 | 0.02 | .64 |
| Sex: Male | 0.45 | 0.13 | <.001* | 0.00 | 0.02 | .87 | -0.03 | 0.02 | .29 |
| BMI | 0.05 | 0.02 | .01* | 0.00 | 0.00 | .31 | 0.00 | 0.00 | .64 |
| FPI | 0.07 | 0.03 | .05* | 0.00 | 0.00 | .76 | 0.00 | 0.01 | .79 |
| FMM | 0.08 | 0.07 | .24 | 0.01 | 0.01 | .25 | 0.00 | 0.01 | .97 |
| FPI * FMM | -0.04 | 0.01 | .008* | 0.00 | 0.00 | .84 | 0.00 | 0.01 | .78 |
|  | Adj. R<sup>2</sup> = 0.19 |  |  | Adj. R<sup>2</sup> = -0.03 |  |  | Adj. R<sup>2</sup> = -0.03 |  |  |
|  | F<sub>(8,71)</sub> = 3.37 |  |  | F<sub>(8,71)</sub> = 0.71 |  |  | F<sub>(8,70)</sub> = 0.69 |  |  |
|  | p = .002* |  |  | p = .69 |  |  | p = .70 |  |  |
| * Statistically significant; SE = standard error; Adj. = adjusted; LAS: Lateral Ankle Sprain; CAI = Chronic Ankle Instability; BMI = Body Mass Index; FPI = Foot Posture Index |  |  |  |  |  |  |  |  |  |

**Table 4.**
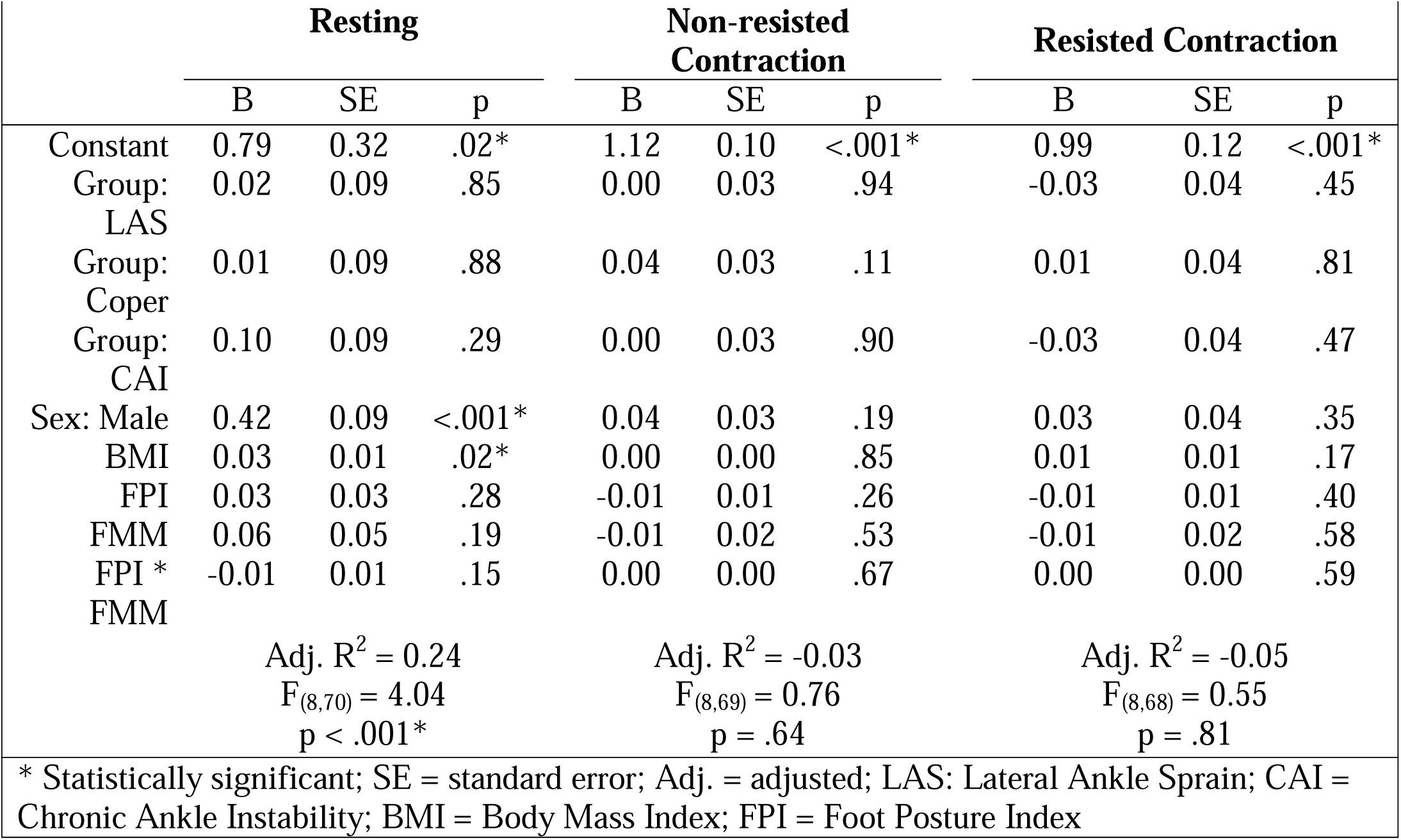
Results of the multiple linear regression analysis predicting ultrasound measures of flexor digitorum brevis resting size and during motor activation

**Table 5.**
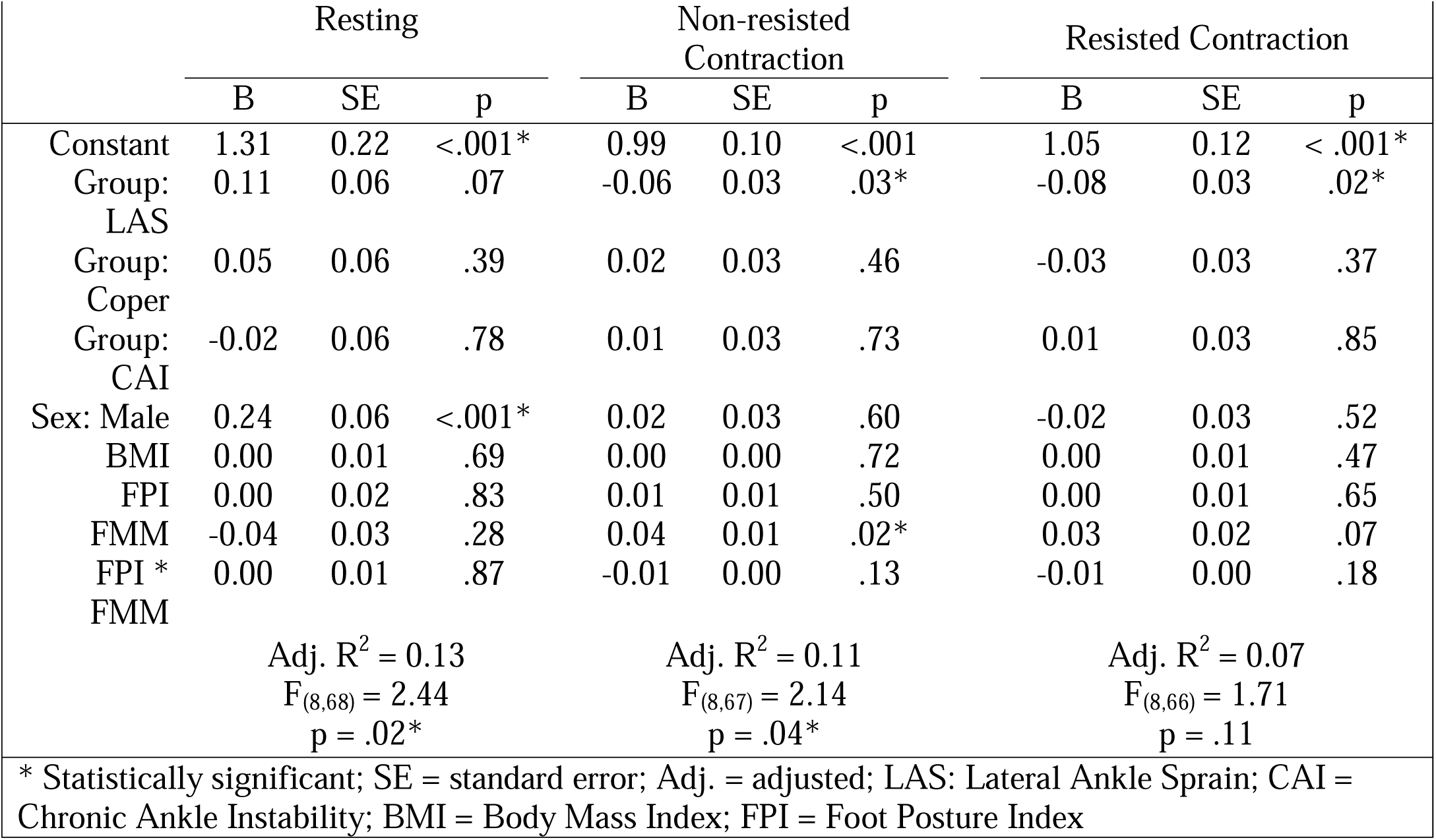
Results of the multiple linear regression analysis predicting ultrasound measures of quadratus plantae resting size and during motor activation

**Table 6.**
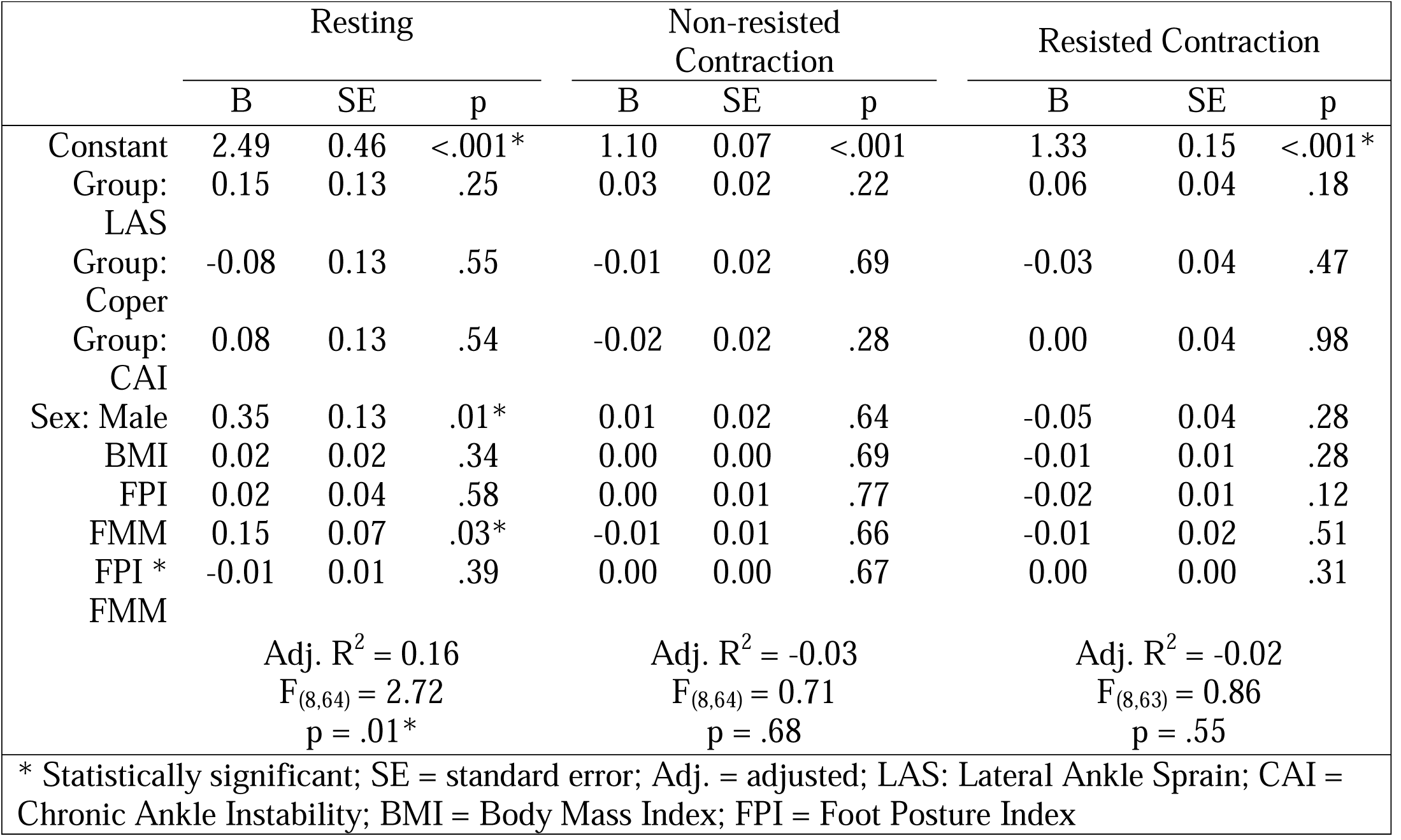
results of the multiple linear regression analysis predicting ultrasound measures of flexor hallucis brevis resting size and during motor activation

## DISCUSSION

This study assessed the effects of ankle injury status on IFM resting size and activation during contractions, while factoring sex, body mass, and foot phenotype, using innovative and clinically accessible ultrasound measures. The primary finding was that there were significant group differences in QP activation, with the CAI group demonstrating deficits in both non-resisted and resisted contraction conditions. The Coper group demonstrated deficits in the non-resisted QP AR measure only. Sex was associated with resting IFM size and was a significant predictor for all muscles in the regression models.

### Resting Measures

In the assessment of injury history on resting IFM cross-sectional area measurements, group was not a significant factor in this study. This was somewhat unexpected as Feger and colleagues^10^ reported large decreased adductor hallucis obliquus and FHB muscle volume in individuals with CAI compared to healthy controls. It is plausible that methodological differences in how the IFM were measured is, in part, responsible for the disparity. Feger and colleagues^10^ used MRI image slices to reconstruct a three-dimensional image of each IFM used to calculate a volumetric measure. The FHB has a medial and lateral head, so it is plausible that MRI is better suited to capture volumetric loss in geometrically complex muscles. It is also plausible that disparity in self-reported ankle-foot function was also a factor. The participants in the study conducted by Feger and colleagues^10^ reported substantially higher disability on the FAAM. It is plausible that the participants included in our study had less neuromotor impairment compared to the sample studied by Feger and colleagues.^10^ Since LAS and CAI are incurred by a diverse population and are highly heterogenous conditions as it pertains to scope and severity of impairment,^4^ it is possible that there are other important covariates such as sex, body mass, and foot phenotype^9,13^ that have not been previously considered.

Sex had fair associations with all resting IFM measures. These results are consistent with those reported by Abe and colleagues^9^ who assessed the relationships of IFM cross-sectional measures to toe strength. They found that men had significantly larger AbdH and FDB muscles compared to women.^9^ It is highly plausible that sex hormones, such as androgens, may also influence the size of the IFM. Males have higher testosterone levels, a primary androgen hormone, which is a primary factor for sex-related related disparities in muscle mass.^30^

Both sex and foot phenotype were significant predictors of AbdH and FHB muscle size. Foot phenotype also contributed to resting FHB measures, with fair associations observed. It is plausible that these findings are related to sex-related differences in foot structure and function. Fukano and colleagues^31^ found that females had greater foot flexibility and deformation under load. Additionally, greater foot deformation has been associated with participants with higher BMIs.^32^ BMI was found to be a significant predictor for both AbdH and FDB muscle cross-sectional area, two muscles that have an important role in support of foot architecture. Therefore, it is important to consider the covariates of sex, body mass, and foot phenotype in evaluation of resting IFM size following ankle-foot injury.

#### Contraction Measures

Within groups, the participants demonstrated increased activation ratios in all IFMs assessed with resistance. This finding was expected and likely reflects increased motor unit recruitment required to counter the external moment. These findings are similar to the increased AR measured in the fibularis and gluteal muscle groups in the same participants.^27^

In the unadjusted analysis, both Coper and CAI groups demonstrated moderate, significant decreases in non-resisted QP AR compared to controls. Furthermore, the CAI group demonstrated large, significantly decreased resisted ARs in the QP compared to both LAS and control groups. This suggests that while individuals in the Coper group were able to activate their QP muscle when resistance was applied, the CAI group demonstrated impaired activation compared to controls. As task difficulty increased, the mean difference in motor activation between CAI and control groups also increased. This is especially interesting considering the function of the QP. In addition to assisting in flexing the toes, the QP neutralizes the adduction moment created by the flexor digitorum longus and contributes to foot pronation.^33^ It is plausible that neuromotor impairment of the QP contributes, in part, to the inverted foot and externally rotated shank commonly observed in this clinical group.^34^ When factored with the other covariates, both LAS group and foot phenotype were significant predictors for QF activation. It is plausible that tibial neuromotor impairment that affects a proportion of grade II and III LASs^7,8^ may be contributory to these findings.

Another study employed USI to compare gluteal muscle thickness and activation during side-lying table top abduction and band-walking exercises in the same participants.^27^ One of the primary observations was that the gluteus medius AR in CAI group was significantly smaller contrasted to Copers, LAS, and healthy controls as task difficulty increased.^27^ In addition, neuromotor compensations were observed, as the CAI group relied more on the gluteus maximus muscle to complete the exercise.^27^ The authors suggested this may be due to central motor alterations that occur after injury that may affect the motor recruitment in the area. ^27^ It is plausible that the same phenomenon was observed in QP muscle of these individuals.

There was a small inverse correlation observed between resisted FHB AR and hallux and lesser toe strength, with greater activation observed in individuals with lower force output. It is plausible that altered motor activation, with greater contribution from the IFM, is required to compensate for weaker flexor hallucis longus and flexor digitorum longus strength. Conversely, it equally plausible that individuals with greater toe flexion output are more efficient and require less summation of synergistic motor units. Lastly, translation of the muscle belly away from the assessment site or compression by neighboring muscles during strong contraction may have artificially gave the appearance of decreased activation. Further investigation into these suppositions is warranted.

Sex was not associated with IFM activation in both non-resisted and resisted conditions. Assessment of IFM ARs are not a measure of strength, but a measure of neuromotor activation. It is possible that females and males utilize the same amount of muscle activation to perform the task, even though sex-related differences in resting IFM muscle sizes were observed.

### Clinical Implications

These findings, when synthesized together, can be contextualized using the theoretical framework postulated by McKeon and colleagues.^35^ In the “foot core” paradigm, there is an interplay of active and passive foot subsystems, with the active subsystem represented by the IFM and the passive subsystem provided by the multitude of joint capsules and ligamentous in the foot. The external moments imposed by body mass during foot deformation, which is modulated by sex-related joint laxity, is resisted by the IFM. Since single-point resting measures provide of IFM cross-sectional measures insight to associated atrophy or hypertrophy, it appears that resting measures are more a function of foot phenotype and external loads rather than injury history. The third facet of the “foot core” triad presented by McKeon and colleagues^35^ is the neural subsystem. Clinical group was a significant factor for QP neuromotor function, represented by non-resisted and resisted activation.

Based on the findings of this study, routine examination of the IFM at rest and during activation using USI following LAS and CAI cannot be recommended at this time. However, clinicians may consider using these measures as a clinical correlate if the patient is suspected of having neuromotor impairment following injury (i.e. tarsal tunnel neuropraxia).

### Limitations and Strengths

This study was not without limitations. The authors employed a cross-sectional design in the assessment of clinical group on resting IFM size and motor activation. As a result, causation cannot be implied. The sample consisted of young adult, nulliparous women and men with and without LAS and CAI, which limits external validity to similar populations. Since ankle-foot impairment varies in scope and depth following LAS and in CAI, heterogeneity of clinical presentation is likely. This study also had strengths worth mentioning. This is the first study to assess IFM size and neuromotor function and identify findings using a clinically accessible imaging modality. This study also considered multiple contributory factors during analysis, which provides greater insight to the clinical outcomes employed in this investigation.

## CONCLUSION

IFM resting ultrasound measures were primarily determined by sex, BMI, and foot phenotype and not injury status. Group differences in contraction measures of the QP were observed in unadjusted and adjusted analyses, explaining a small proportion of the variance. The clinical utility of these IFM ultrasonographic assessments in young adults with LAS and CAI is limited. Based on the findings of this study, routine examination of the IFM at rest and during activation using USI following LAS and CAI cannot be recommended at this time. However, clinicians may consider using these measures as a clinical correlate if the patient is suspected of having neuromotor impairment following injury.

## Data Availability

The data that support the findings of this study are available from the corresponding author, JJF, upon reasonable request.

## Acknowledgements

Abbis H Jaffri, PT, PhD for his assistance during examinations. Clair Seckner and Mary Cowden for their assistance in measurement of ultrasound images.

## REFERENCES

1. Waterman BR. The epidemiology of ankle sprains in the united states. J Bone Jt Surg Am. 2010;92(13):2279–2284. doi:10.2106/JBJS.I.01537

2. Wikstrom EA, Brown CN. Minimum Reporting Standards for Copers in Chronic Ankle Instability Research. Sports Med. 2014;44(2):251–268. doi:10.1007/s40279-013-0111-4

3. Doherty C, Bleakley C, Hertel J, Caulfield B, Ryan J, Delahunt E. Recovery from a first-time lateral ankle sprain and the predictors of chronic ankle instability: a prospective cohort analysis. Am J Sports Med. 2016;44(4):995–1003. doi:10.1177/0363546516628870

4. Hertel J, Corbett RO. An Updated Model of Chronic Ankle Instability. J Athl Train. 2019;54(6):572–588. doi:10.4085/1062-6050-344-18

5. Fraser JJ, Koldenhoven RM, Jaffri AH, et al. Foot impairments contribute to functional limitation in individuals with ankle sprain and chronic ankle instability. Knee Surg Sports Traumatol Arthrosc. July 2018. doi:10.1007/s00167-018-5028-x

6. Fraser JJ, Hertel J. Joint Mobility & Stability Strategies for the Ankle. In: 29.2 Neurology in Orthopaedics. Alexandria, VA: Academy of Orthopaedic Physical Therapy, APTA, Inc.; 2019. https://doi.org/10.17832/ISC.2019.29.2.5.

7. Nitz AJ, Dobner JJ, Kersey D. Nerve injury and grades II and III ankle sprains. Am J Sports Med. 1985;13(3):177–182.

8. Jazayeri Shooshtari SM, Didehdar D, Moghtaderi Esfahani AR. Tibial and peroneal nerve conduction studies in ankle sprain. Electromyogr Clin Neurophysiol. 2007;47(6):301–304.

9. Abe T, Tayashiki K, Nakatani M, Watanabe H. Relationships of ultrasound measures of intrinsic foot muscle cross-sectional area and muscle volume with maximum toe flexor muscle strength and physical performance in young adults. J Phys Ther Sci. 2016;28(1):14–19. doi:10.1589/jpts.28.14

10. Feger MA, Snell S, Handsfield GG, et al. Diminished Foot and Ankle Muscle Volumes in Young Adults With Chronic Ankle Instability. Orthop J Sports Med. 2016;4(6). doi:10.1177/2325967116653719

11. Fraser JJ, Mangum LC, Hertel J. Test-retest reliability of ultrasound measures of intrinsic foot motor function. Phys Ther Sport. 2018;30:39–47. doi:10.1016/j.ptsp.2017.11.032

12. Fraser JJ, Hertel J. Effects of a 4-Week Intrinsic Foot Muscle Exercise Program on Motor Function: A Preliminary Randomized Control Trial. J Sport Rehabil. 2019;28(4):339–349. doi:10.1123/jsr.2017-0150

13. Zhang X, Aeles J, Vanwanseele B. Comparison of foot muscle morphology and foot kinematics between recreational runners with normal feet and with asymptomatic over-pronated feet. Gait Posture. 2017;54:290–294. doi:10.1016/j.gaitpost.2017.03.030

14. Gribble PA, Delahunt E, Bleakley C, et al. Selection criteria for patients with chronic ankle instability in controlled research: a position statement of the International Ankle Consortium. Br J Sports Med. 2014;48(13):1014–1018. doi:10.1136/bjsports-2013-093175

15. Martin RL, Irrgang JJ, Burdett RG, Conti SF, Van Swearingen JM. Evidence of validity for the foot and ankle ability measure (FAAM). Foot Ankle Int. 2005;26(11):968–983.

16. Carcia CR, Martin RL, Drouin JM. Validity of the Foot and Ankle Ability Measure in Athletes With Chronic Ankle Instability. J Athl Train. 2008;43(2):179–183.

17. Donahue M, Simon J, Docherty CL. Reliability and validity of a new questionnaire created to establish the presence of functional ankle instability: the IdFAI. Athl Train Sports Health Care. 2013;5(1):38–43. doi:10.3928/19425864-20121212-02

18. Hays RD, Bjorner JB, Revicki DA, Spritzer KL, Cella D. Development of physical and mental health summary scores from the patient-reported outcomes measurement information system (PROMIS) global items. Qual Life Res. 2009;18(7):873–880. doi:10.1007/s11136-009-9496-9

19. Woby SR, Roach NK, Urmston M, Watson PJ. Psychometric properties of the TSK-11: A shortened version of the Tampa Scale for Kinesiophobia: Pain. 2005;117(1):137–144. doi:10.1016/j.pain.2005.05.029

20. Godin G, Shephard RJ. Godin leisure-time exercise questionnaire. Med Sci Sports Exerc. 1997;29(6):36–38.

21. Revicki DA, Kawata AK, Harnam N, Chen W-H, Hays RD, Cella D. Predicting EuroQol (EQ-5D) scores from the patient-reported outcomes measurement information system (PROMIS) global items and domain item banks in a United States sample. Qual Life Res. 2009;18(6):783–791. doi:10.1007/s11136-009-9489-8

22. Redmond AC, Crosbie J, Ouvrier RA. Development and validation of a novel rating system for scoring standing foot posture: The Foot Posture Index. Clin Biomech. 2006;21(1):89–98. doi:10.1016/j.clinbiomech.2005.08.002

23. Williams DS, McClay IS. Measurements Used to Characterize the Foot and the Medial Longitudinal Arch: Reliability and Validity. Phys Ther. 2000;80(9):864–871.

24. McPoil TG, Vicenzino B, Cornwall MW, Collins N, Warren M. Reliability and normative values for the foot mobility magnitude: a composite measure of vertical and medial-lateral mobility of the midfoot. J Foot Ankle Res. 2009;2(1):6. doi:10.1186/1757-1146-2-6

25. Fraser JJ, Koldenhoven R, Hertel J. Reliability of measures of ankle-foot morphology, mobility, and strength. Int J Sports Phys Ther. 2017;12(7):1134–1149. doi:10.16603/ijspt20171134

26. Hashimoto T, Sakuraba K. Assessment of Effective Ankle Joint Positioning in Strength Training for Intrinsic Foot Flexor Muscles: A Comparison of Intrinsic Foot Flexor Muscle Activity in a Position Intermediate to Plantar and Dorsiflexion with that in Maximum Plantar Flexion Using Needle Electromyography. J Phys Ther Sci. 2014;26(3):451–454. doi:10.1589/jpts.26.451

27. Koldenhoven RM, Fraser JJ, Saliba SA, Hertel J. Ultrasonography of Gluteal and Fibularis Muscles During Exercises in Individuals With A History of Lateral Ankle Sprain. J Athl Train. October 2019. doi:10.4085/1062-6050-406-18

28. Cohen J. Statistical Power Analysis for the Behavioral Sciences. Academic Press; 2013.

29. Portney LG, Watkins MP. Foundations of Clinical Research: Applications to Practice. Pearson/Prentice Hall; 2009.

30. Mooradian AD, Morley JE, Korenman SG. Biological actions of androgens. Endocr Rev. 1987;8(1):1–28. doi:10.1210/edrv-8-1-1

31. Fukano M, Fukubayashi T. Gender-based differences in the functional deformation of the foot longitudinal arch. The Foot. 2012;22(1):6–9. doi:10.1016/j.foot.2011.08.002

32. Fritz B, Schmeltzpfenning T, Plank C, Hein T, Grau S. Anthropometric influences on dynamic foot shape: Measurements of plantar three-dimensional foot deformation. Footwear Sci. 2013;5(2):121–129. doi:10.1080/19424280.2013.789559

33. Pretterklieber B. Morphological characteristics and variations of the human quadratus plantae muscle. Ann Anat - Anat Anz. 2018;216:9–22. doi:10.1016/j.aanat.2017.10.006

34. Moisan G, Descarreaux M, Cantin V. Effects of chronic ankle instability on kinetics, kinematics and muscle activity during walking and running: A systematic review. Gait Posture. 2017;52:381–399. doi:10.1016/j.gaitpost.2016.11.037

35. McKeon PO, Hertel J, Bramble D, Davis I. The foot core system: a new paradigm for understanding intrinsic foot muscle function. Br J Sports Med. 2015;49(5):290–290. doi:10.1136/bjsports-2013-092690

